# GaitEncoder: A Foundation Model of Gait Kinematics for Diverse Clinical Applications and Pathologies

**DOI:** 10.64898/2026.07.07.26357479

**Authors:** R. Daniel Magruder, Selim Gilon, Antoine Falisse, Scott D. Uhlrich

## Abstract

Quantitative gait analysis could enhance personalized treatment for many movement-related conditions; however, it is not routinely integrated into clinical care. Advances in mobile sensing, such as smartphone-based motion capture, enable rapid clinical gait assessment, but extracting actionable insights remains challenging. Although machine learning models can support clinical decisions from gait data, they typically require costly task- and condition-specific datasets, which limits progress across various gait-related conditions. Here we present a generative foundation model of walking kinematics that enables various downstream clinical tasks across diverse patient populations using clinically accessible smartphone video–based gait analysis. We aggregated eight gait datasets comprising 657 individuals across seven unique pathologies. Using weakly-supervised learning, we trained a variational autoencoder to distill high-dimensional gait kinematics into a 16-dimensional learned latent representation. We demonstrate generalizability across four downstream clinical tasks spanning pathologies both seen and unseen during training, with and without model fine-tuning, including: 1) classification of neuromuscular disorders unseen during training, 2) predicting clinical severity scores for individuals with Parkinson’s disease, 3) tracking of subacute recovery post-stroke, and 4) generating patient-specific kinematic changes following total hip arthroplasty. Our model also computes a deviation from mean unimpaired (DMU) score, an interpretable scalar metric that captures an individual’s deviation from typical unimpaired gait, providing rapid, holistic quantification of impairment. This generalizable model provides a foundation for clinically actionable tools that translate mobile sensing–derived gait data into precise biomechanical insights for clinical research and decision-making. The open-source model is deployed in the cloud for automated smartphone video–based gait analysis on our freely available OpenCap platform.

## 1 Introduction

Human gait kinematics can enable precise disease severity monitoring, personalized treatment selection, and tracking of rehabilitation progress. For example, in patients with cerebral palsy, the Gait Deviation Index ^1^ is a scalar metric that can be used to monitor disease progression and to predict and track treatment effects ^2,3^. More complex representations of gait can predict kinematic outcomes across multiple potential surgeries and suggest alternative interventions, such as strength training, prior to a final treatment decision ^4^. As another example, for patients with knee osteoarthritis, gait analysis enables patient-specific selection of joint-offloading interventions that reduces pain ^5,6^. Furthermore, pre-operative gait parameters can predict which individuals are likely to respond well to knee replacement surgery ^7^. Unfortunately, these insights traditionally require specialized gait labs, which have seen limited clinical adoption. Thus, treatment of most movement-related conditions is typically not informed by in-depth gait analysis due to the cost, time, and expertise required to collect and interpret the data ^8–10^.

Recent advances in mobile sensing enable rapid clinical gait analysis using low-cost hardware ^11–14^. For example, we developed OpenCap, an open-source platform that can measure 3D skeletal motion in minutes in the clinic using one or more smartphone videos ^15–18^. Despite the growing accessibility, translating the resulting gait kinematics into clinical insights remains a fundamental barrier. A single walking trial produces complex time series kinematic data across dozens of joints. Thus, gait is often reduced to scalar summary metrics ^19^ (e.g., gait speed, step length, cadence) that are interpretable but discard the majority of the signal. Full kinematic analysis can support clinical decision making; however, it is largely limited to specialists with sufficient time and expertise required for analysis ^4^. There is a need for a method that can distill high-dimensional gait data into features that are both clinically meaningful and actionable.

Machine learning approaches are a powerful way to generate actionable insights from gait data ^3,20,21^. However, they often require large, condition-specific datasets (e.g., n=933 children with cerebral palsy ^22^), involve hand-engineering of input features ^23^, and are constrained to specific downstream clinical tasks ^24^. Thus, there remains a need for models that generalize across movement-related conditions and downstream prediction tasks. This would be especially transformative for rare movement-related conditions where large gait datasets do not exist and would be difficult to obtain.

Foundation models are a promising approach to this problem, as they can synthesize a variety of inputs and perform multiple downstream tasks. Cancer detection has greatly benefited from the development of foundation imaging models with a single model evaluating biomarkers, identifying cells, and detecting common and rare cancers at clinically relevant accuracy and specificities ^25^. Furthermore, foundation models in medical imaging readily scale to novel tasks with minimal data ^26^. A foundation model of gait kinematics should similarly perform multiple downstream tasks, scale well for many movement conditions within and outside the training dataset, and be able to improve specific downstream task performance (e.g., predict therapeutic effects) through fine-tuning on small datasets. Training these foundation models requires large, diverse datasets; however, there is a lack of sufficiently large, pathologically diverse, and biomechanically accurate gait datasets.

Self-supervised learning is a common method to train foundation models when data labeling is difficult or scarce. It also enables training on larger, more diverse datasets ^27^, as demonstrated in previous work applied to gait kinematics ^28^ and kinetics ^29^. Variational autoencoders (VAE) are generative models, conducive to self-supervised learning. VAEs comprise an encoder network that distills input data into a representative latent (low-dimensional) feature space and a decoder network that reconstructs the original input from the latent space ^30–32^. VAEs have been effective in pathology-specific gait feature reduction ^33,34^ and in tokenizing human movement for downstream learning tasks ^35^. While self-supervision is appealing because it avoids explicit clinical labels, functional measures such as gait speed—though coarse—may provide weak supervision that guides training toward clinical interpretability. A VAE trained on a large, pathologically diverse dataset and weakly guided by such functional measures could capture fundamental features of diverse movement conditions while providing clinically useful insights for both seen and unseen gait pathologies.

The purpose of our study was to develop and evaluate a generative foundation model that learns a generalizable representation of gait kinematics to support a range of downstream clinical tasks. We assessed our model’s ability to 1) reconstruct salient kinematic features characteristic of various gait-related pathologies, 2) perform multiple downstream clinical tasks with and without model fine-tuning, and 3) generalize across a variety of pathologies both within and outside the training set. To demonstrate this, we evaluated our model on four clinically relevant tasks: identifying rare neuromuscular diseases from controls; quantifying motor impairment in agreement with standard clinician- and self-reported scales; tracking patient recovery during rehabilitation; and predicting the effect of surgery on gait. Importantly, these tasks span conditions that were both in training (post-stroke and hip osteoarthritis) and those entirely withheld (two neuromuscular diseases and Parkinson’s disease), and they include kinematics derived from both markered and smartphone-based markerless motion capture. The purpose of these diverse evaluations is to demonstrate the generalizability of our model, its utility for zero-shot prediction, and the ability to fine-tune it with small population- and task-specific datasets. To facilitate widespread use of our open-source model, we provide a cloud-based tool for automatically extracting our gait features from marker-based motion capture or OpenCap smartphone video–based kinematic data.

## 2 Results

Here we present GaitEncoder, a VAE that reduces gait kinematics (32 joints over 24 time points) into a 16-dimensional latent representation (Figure 1). We aggregated a dataset (n=657) comprising 7 gait-related pathologies and trained our final model on this entire dataset. For evaluations in this paper, we trained the VAE on a subset of the dataset (n=381; 4 pathologies), enabling evaluation on three unseen pathologies (Figure 2a). We found 16 features sufficient to encode salient features of gait kinematics with low kinematic reconstruction error (mean absolute error [MAE]=3.5° across all joints, Figure S1). Three movement-related conditions were held out of training entirely: myotonic dystrophy, facioscapulohumeral muscular dystrophy (FSHD), and Parkinson’s disease. Their reconstruction MAE were 4.8°, 4.3°, and 5.1°, respectively. The held-out Parkinson’s disease dataset (n=26) comprised kinematics derived from marker-based motion capture, while the myotonic dystrophy (n=55) and FSHD (n=26) datasets comprised kinematics from smartphone videos (OpenCap). From this latent representation, we derived the deviation from mean unimpaired (DMU) score as a scalar summary of gait impairment, where higher values indicate greater deviation from an unimpaired cohort. We used the DMU score and the 16-dimensonal latent representation to perform four downstream clinical tasks. Within these tasks, we tested our latent representation in three modes: zero-shot, fine-tuned, and as a low-dimensional input to a separate downstream predictor network.

**Fig. 1.**
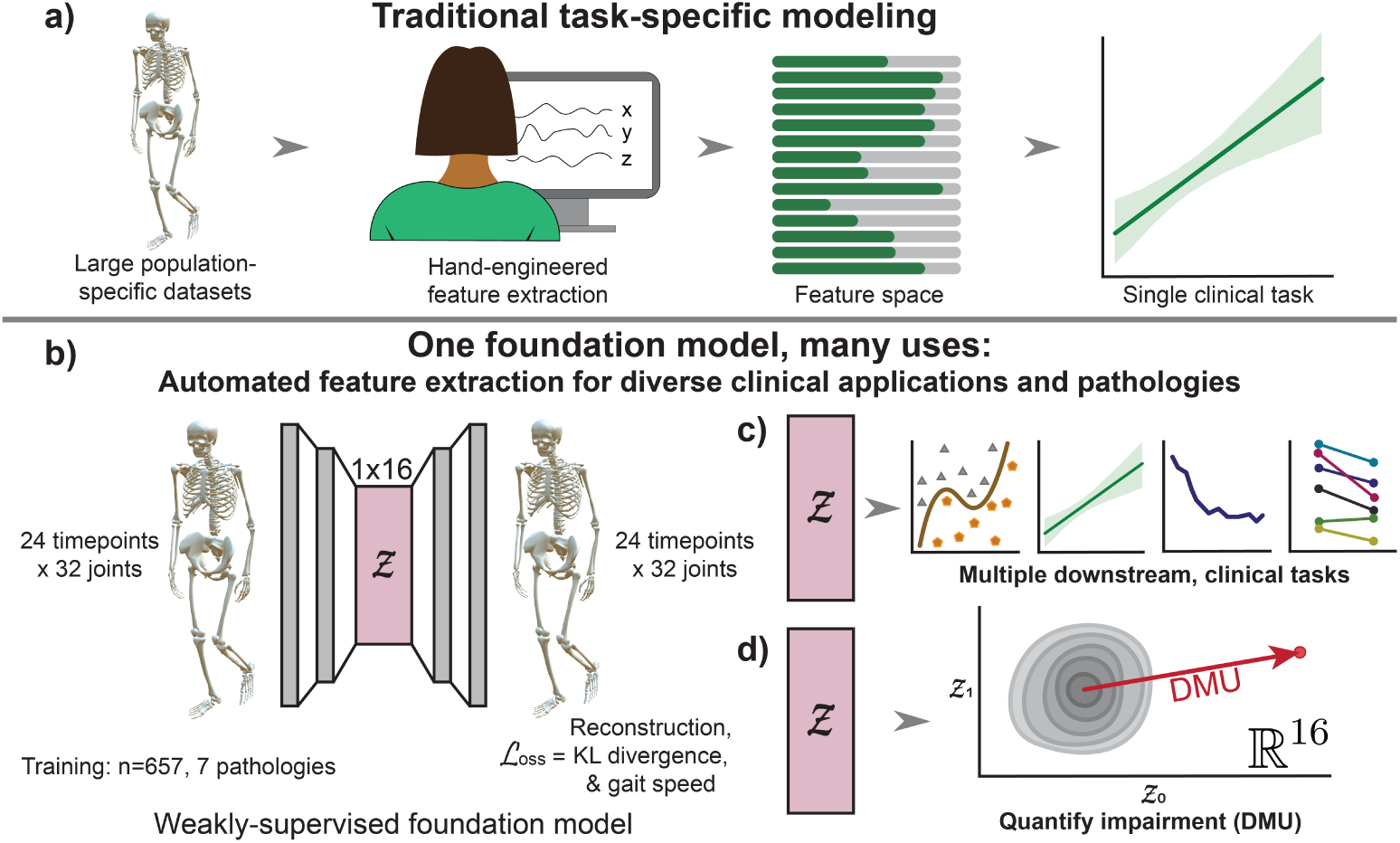
A foundation model for gait analysis. Our foundation model automates gait kinematic feature extraction, enabling a variety of downstream clinical tasks. **a)** Predictive modeling using gait data has traditionally required an expert to hand-engineer features and train a model for a single downstream task (e.g., one population and outcome measure). This approach is time-consuming and requires large datasets. **b)** Our automated feature extraction method uses a variational autoencoder and provides a 16-feature latent space (*z*) that encodes whole-body kinematics. **c)** We demonstrate that the latent representations can be used in a variety of clinically relevant tasks (i.e., disease classification, severity quantification, rehabilitation monitoring, and surgical outcome prediction). **d)** The 16-feature (R16) latent representations can be further distilled to a single score representing holistic gait impairment. This deviation from mean unimpaired (DMU) score quantifies an individual’s deviation from an unimpaired cohort’s latent representation. DMU may be useful for rapid assessments of impairment, while the 16-feature latent space enables more complex and predictive tasks.

**Fig. 2.**
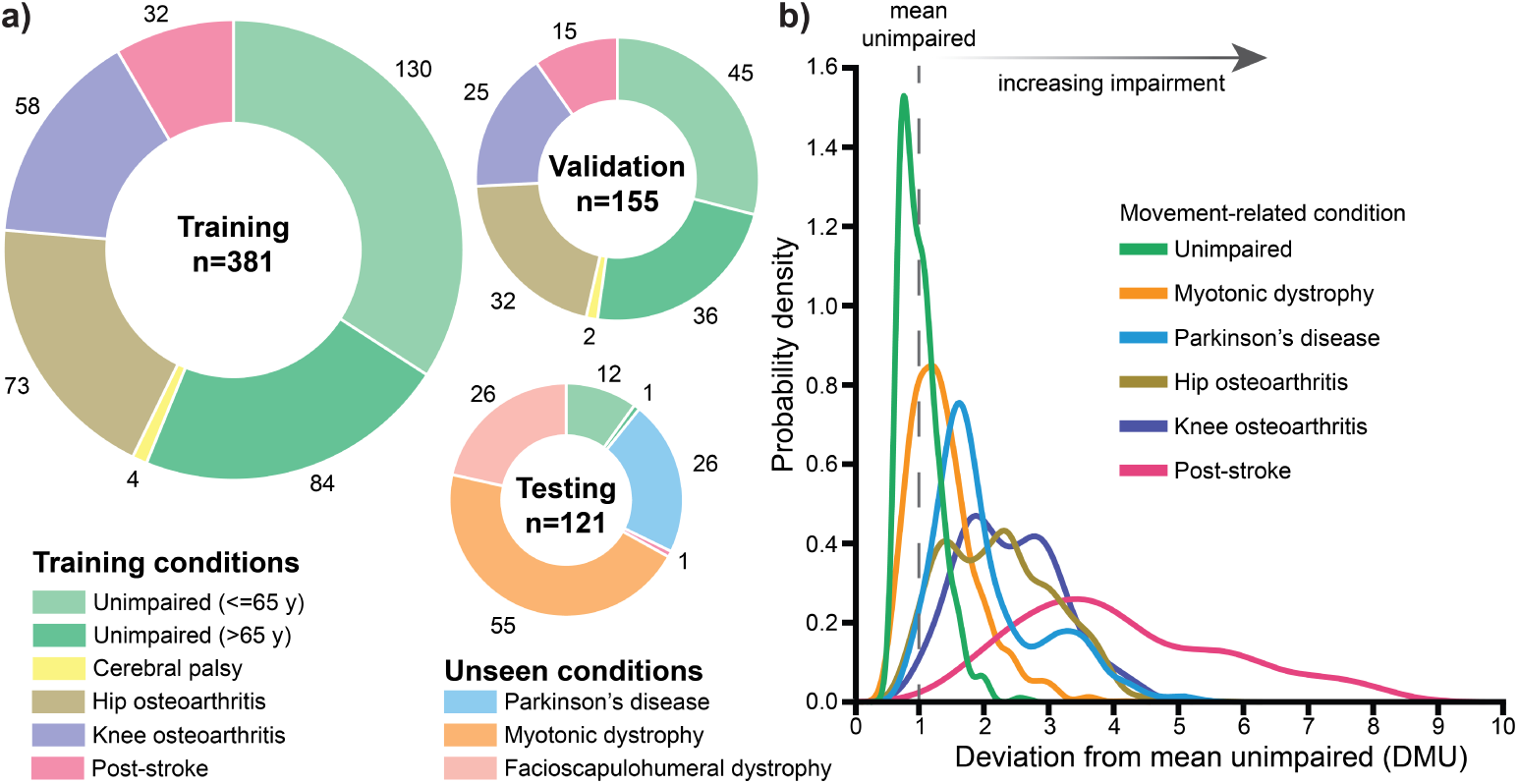
GaitEncoder was trained and evaluated on a kinematically and pathologically diverse dataset. **a)** Four distinct movement pathologies were included in training and validation sets. Individuals with Parkinson’s disease, myotonic dystrophy, and facioscapulohumeral muscular dystrophy were withheld entirely during training. **b)** The scalar-valued deviation from mean unimpaired (DMU) score captures gait impairment across various pathologies. DMU is calculated relative to a set of high-performing unimpaired individuals in the training set (green), and the shown subset of pathologies include individuals in the validation and held-out test set. The grey dashed line represents the mean of training DMU scores, where scores higher than 1.0 deviate increasingly from the unimpaired cohort (i.e., more likely to be impaired).

### 2.1 Downstream application 1: Identifying rare neuromuscular diseases using gait kinematics

More sensitive and objective functional outcome measures are needed to monitor the progression of neuromuscular diseases and detect the efficacy of novel therapeutics ^36^. The ability to distinguish individuals with neuromuscular diseases from a control cohort using gait kinematics captured via accessible motion capture technology represents a key step toward the development of sensitive digital outcome measures. Specifically, myotonic dystrophy weakens lower-limb muscles (e.g., the ankle plantarflexors and knee extensors), resulting in reduced balance and stride lengths ^17,37,38^. While FSHD often affects upper-extremity muscles initially ^17^, it can also weaken lower-extremity muscles (e.g., ankle dorsiflexors and knee flexors), which can cause a variety of impairments, like foot drop ^39^. However, these gait adaptations for myotonic dystrophy and FSHD can be subtle and variable.

We computed GaitEncoder latent representations from previously collected Open-Cap markerless gait kinematics for cohorts with myotonic dystrophy (n=55), FSHD (n=26), and held-out control (n=13) participants. We trained support vector machines for case–control classification, using the computed latent representation as input. The model achieved a class-balanced accuracy of 0.68 for myotonic dystrophy vs. control, despite myotonic dystrophy being withheld during training (i.e., zero-shot). Fine-tuning the GaitEncoder on data from individuals with myotonic dystrophy improved accuracy to 0.81 (evaluated on held-out data). For comparison, models using clinician-informed, hand-engineered features ^17^ for myotonic dystrophy achieved accuracies of 0.53 using gait features and 0.75 using features from nine activities (Figure 3a). Both the zero-shot and fine-tuned GaitEncoder outperformed the model using hand-engineered gait features (McNemar’s test, p<.001). The same pattern was observed for classifying individuals with FSHD (Figure 3b); however, the accuracy difference between the fine-tuned GaitEncoder model (accuracy=0.75) and the model using hand-engineered gait features (accuracy=0.44) did not reach significance (p=0.066). Zero-shot and fine-tuned models were similar in accuracy with the clinician-informed features from nine activities (accuracy=0.67). Notably, the FSHD and myotonic dystrophy cohorts comprise individuals with a wide range of symptoms, including those without visible gait impairment. Thus, it is unlikely that perfect accuracy could be achieved on this classification task from walking data alone.

**Fig. 3.**
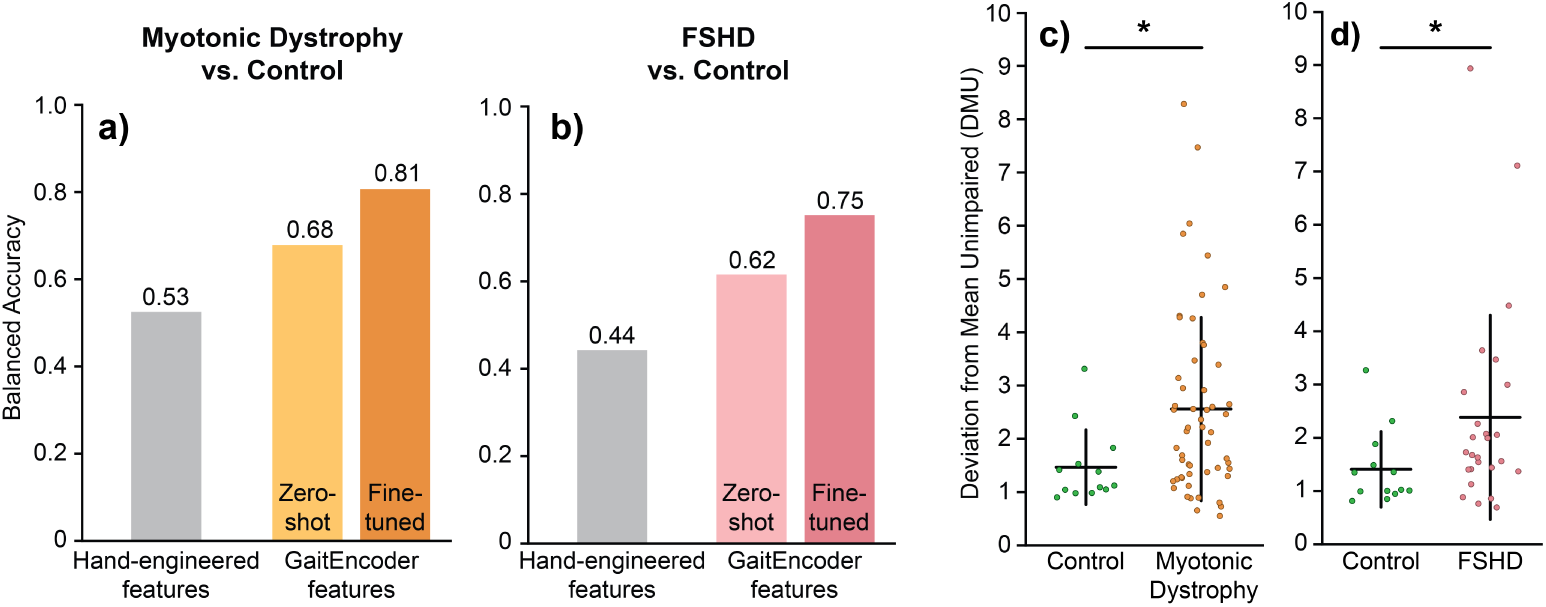
Gait representations identify unseen neuromuscular disorders. GaitEncoder differentiated individuals with and without two rare neuromuscular disorders, myotonic dystrophy and facioscapulohumeral muscular dystrophy (FSHD), which were unseen during model training. **a-b)** We trained classifiers to distinguish individuals with either myotonic dystrophy or FSHD from control. Models that used GaitEncoder’s latent representation (zero-shot, light pink/orange) outperformed models using hand-engineered features of gait (light gray) ^17^. Fine-tuning our GaitEncoder using the myotonic dystrophy or FSHD dataset (dark pink/orange) further improved accuracy. **c-d)** The deviation from mean unimpaired (DMU) score differentiated individuals with each neuromuscular disease from controls. *p*<*0.05.

The scalar DMU score—computed from the zero-shot GaitEncoder latent distribution—was larger on average in the myotonic dystrophy group compared to controls (t=1.91, p=.031, Cohen’s d=.67). We recomputed DMU after fine-tuning the Gait-Encoder on data from the myotonic dystrophy cohort and found further separation between the groups (t=2.23, p=.015, Cohen’s d=.83; Figure 3c). The zero-shot DMU score was not significantly different between FSHD and controls (t=.63, p=.267, Cohen’s d=.22), but was significant after fine-tuning (t=1.766, p=.043, Cohen’s d=.68; Figure 3d). A scalar value from a linear method (single vector decomposition; as used in the Gait Deviation Index ^1^), did not find group differences between myotonic dystrophy (t=.88, p=.810) nor FSHD (t=1.29, p=.897) and the control cohort, highlighting the advantages of the nonlinear VAE model.

### 2.2 Downstream application 2: Predicting patient impairment in agreement with clinician and self-reported scales

Clinician-rated and self-reported measures such as UPDRS-III and UPDRS-II are widely used to assess function in Parkinson’s disease, but they are ordinal, coarse, and difficult to collect remotely ^40^. In contrast, gait kinematics can be measured regularly outside the clinic ^16^, enabling the development of quantitative, pathology-specific impairment scores using models such as GaitEncoder. To support the transition towards more quantitative measures, it is critical to establish construct validity by demonstrating that GaitEncoder captures the same underlying functional impairments reflected in established clinical scales. Here we used linear regression with leave-one-subject-out cross-validation to predict UPDRS scores (Figure 4a) from the GaitEncoder latent representation. GaitEncoder was not trained on data from individuals with Parkinson’s disease. The regression models predicted UPDRS-II with r=.63 (p*<*.001) and UPDRS-III with r=.65 (p*<*.001). While a regressor using the DMU scalar alone could predict UPDRS scores (r=.37–.43, p=.012–.023), it performed worse than the latent-based regressor (Figure 4b). This is unsurprising given the richer kinematic representation contained in the full latent representation. Sub-group analysis of on- and off-medication is reported in the supplement.

**Fig. 4.**
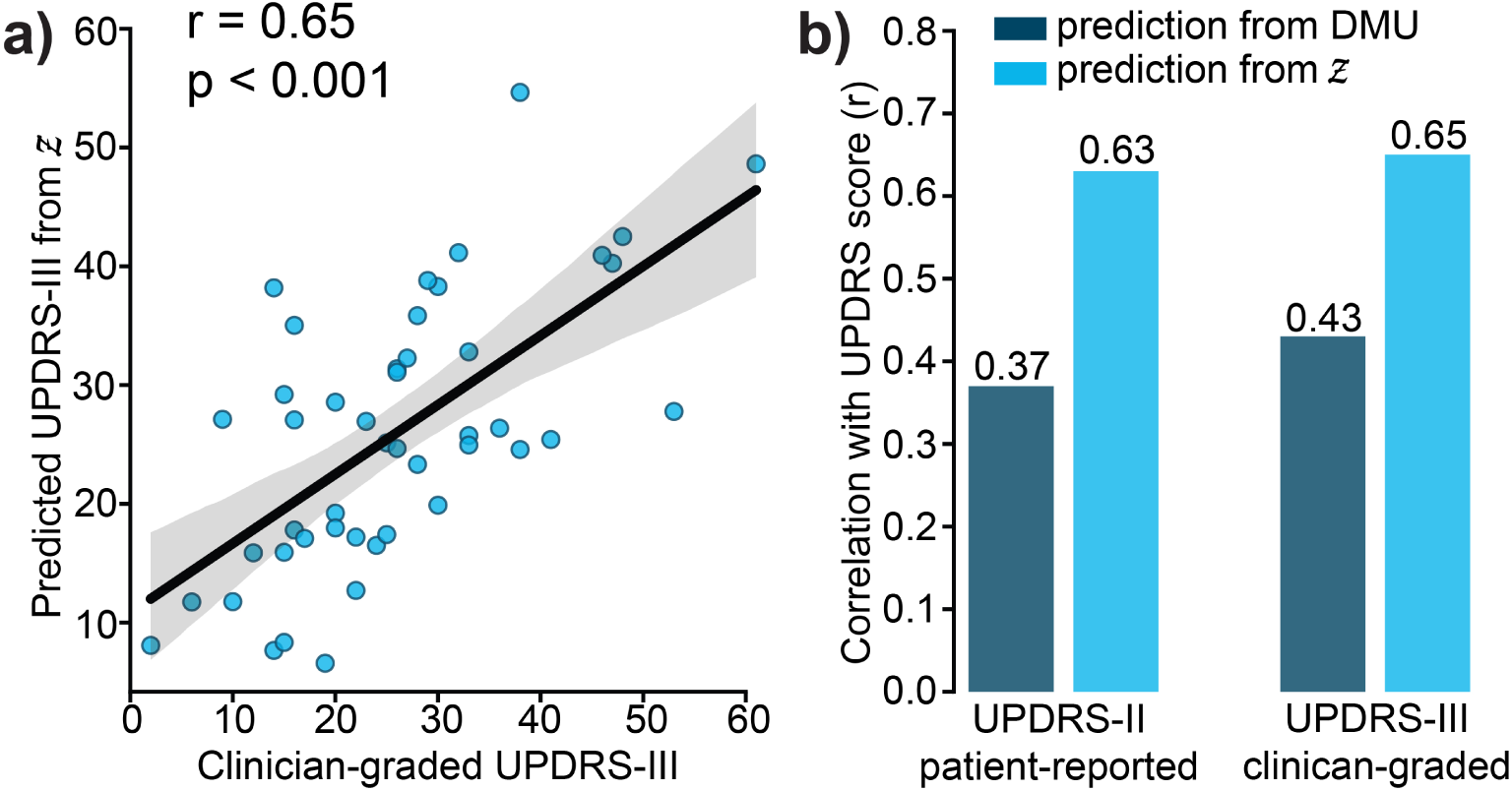
Gait representations predict Parkinson’s disease severity. GaitEncoder predicts patient- and clinician-graded movement scales for Parkinson’s Disease (UPDRS-II and -III, respectively). **a**) The GaitEncoder latent representation (*z*) predicts UPDRS-III using linear regression with leave-one-subject-out cross-validation. **b**) The scalar deviation from mean unimpaired (DMU) score also predicts both UPDRS values, but less accurately than the full latent representation.

### 2.3 Downstream application 3: Tracking patient recovery during subacute post-stroke rehabilitation

For individuals recovering from stroke, the ability to quantify gait impairment and improvement through rehabilitation could be used to create personalized treatment protocols or to justify continued therapy. However, gait impairment varies across individuals, making it difficult to identify a single kinematic feature (e.g., a joint angle) that reflects impairment broadly; a holistic score such as the DMU may better capture heterogeneous gait impairments. We report a case study of one individual (67y) recorded with smartphone video–based motion capture for 16 weeks during the subacute phase of stroke recovery. Data collection began at 5 weeks post-stroke, when she was first able to walk without assistance. We captured walking kinematics at a self-selected speed each week at the individual’s home, and we extracted commonly assessed spatiotemporal features of gait along with the GaitEncoder DMU. The DMU improved during early weeks of rehabilitation, followed by more gradual changes in the later weeks (Figure 5a). This recovery trajectory was similar to other gait metrics, like step length symmetry (Figure 5b) and gait speed (Figure 5c).

**Fig. 5.**
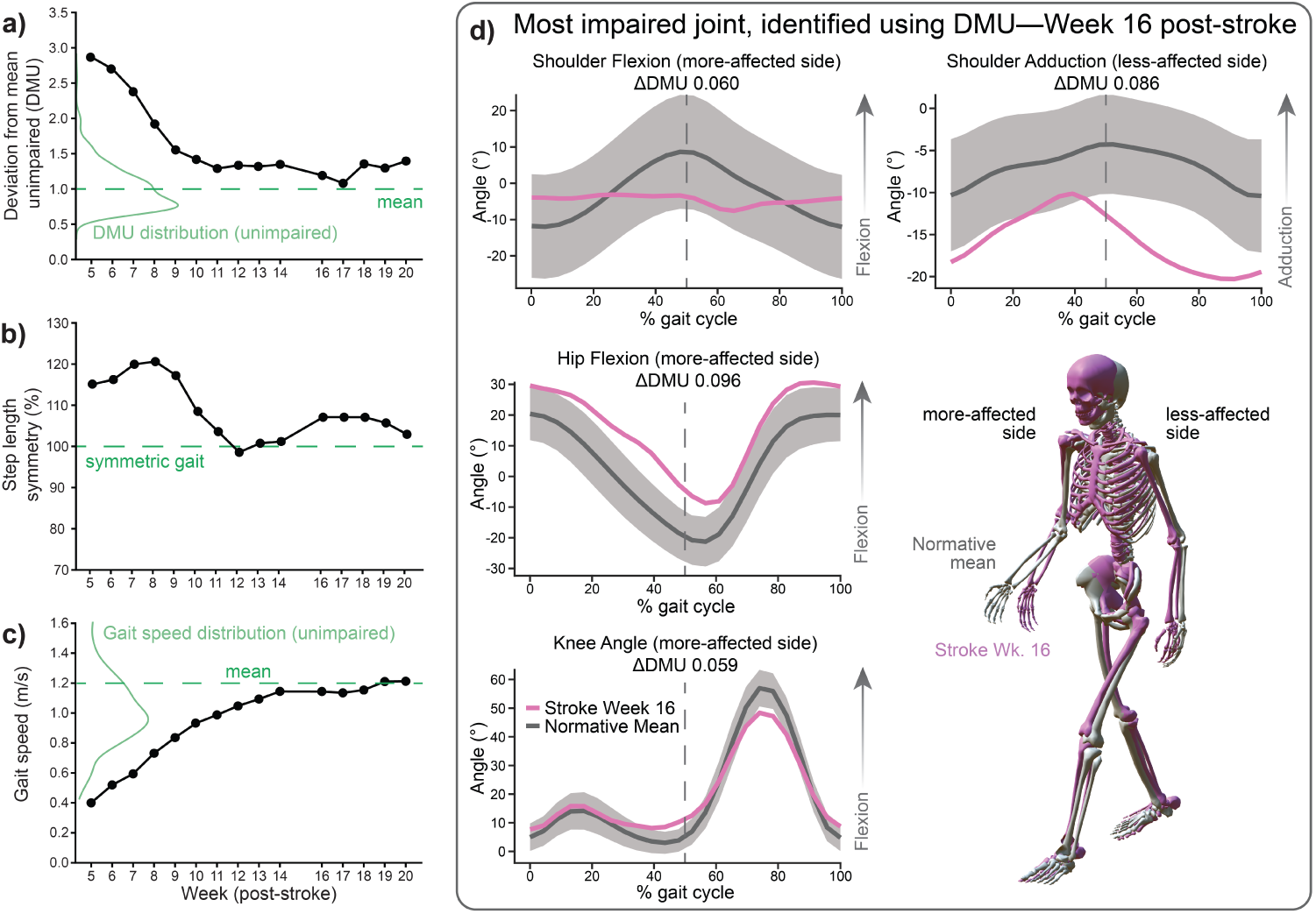
Tracking kinematic impairment during post-stroke rehabilitation. The deviation from mean unimpaired (DMU) score is sensitive to recovery in an individual in the subacute phase after stroke. **a)** DMU improves during early rehabilitation and stabilizes with subtle impairment around week 11 (1.0 corresponds to the unimpaired mean). **b)** Step length symmetry recovers by week 12. **c)** Gait speed increases more gradually than DMU and exceeds the mean unimpaired gait speed at week 19. **d)** To elucidate kinematic differences that persist once gait speed plateaus at week 16, we show the post-stroke participant’s kinematics (pink) vs. average unimpaired kinematics (grey) at 50% of the gait cycle (skeletons). The time series joint kinematics for the four joints that contribute most to the elevated DMU are shown. The change in DMU (ΔDMU) quantifies how replacing each joint trajectory with the unimpaired mean affects the DMU score.

Kinematically, this individual improved rapidly, demonstrating visibly smoother kinematics (Supplemental Videos S1 and S2) over time. However, despite reaching a faster-than-average gait speed by the end of the monitoring period, her DMU remained higher than the normative average of 1.0, indicating a persistent deviation from normative gait. To interpret the elevated DMU, we computed the change in DMU after replacing each joint’s kinematics with normative trajectories; joints producing the largest DMU change contribute most to the elevated DMU. At week 16 when her gait speed plateaued, the joints most contributing to elevated DMU included hip flexion, knee flexion, and arm flexion on the impaired side, as well as arm adduction on the unimpaired side (Figure 5d). We did not perform statistical testing on this case study, but the consistency of the DMU changes with established clinical gait features supports its validity. Moreover, DMU’s ability to identify subtle kinematic deviations that persist after coarser gait metrics (i.e., gait speed) normalize highlights its potential to guide individualized rehabilitation.

### 2.4 Downstream application 4: Predicting kinematic improvements after total hip arthroplasty

A core objective of personalized medicine is to predict the effects of potential interventions based on the characteristics of an individual. We leveraged GaitEncoder’s latent representation to train an intervention-effect prediction model and subsequently used its generative capability to reconstruct post-total hip replacement kinematics from pre-surgical kinematics (n=91 dataset ^41^). Specifically, we trained a feedforward neural network with a single hidden layer (64 nodes) to transform a participant’s presurgical to their post-surgical latent representations (i.e., *surgical effect predictor* in Figure 6a). To determine the capacity of the surgical effect predictor to infer personalized responses to surgery, we compared the reconstructed post-surgical kinematics to the estimated kinematics when applying the average effect of the surgery to all individuals. In the training set, the MAE for post-surgical lower limb kinematics was 4.4±0.8° for our VAE-based surgical effect predictor and 5.7±2.1° for the average effect of surgery. In the held-out validation set, the VAE-based surgical effect predictor predicted post-surgical kinematics with an MAE of 4.7±1.0°, which was lower than that of the average effect of surgery (MAE=5.9±2.5°; paired t-test; t=3.14, p=.004, Cohen’s d=.60, Figure 6b).

**Fig. 6.**
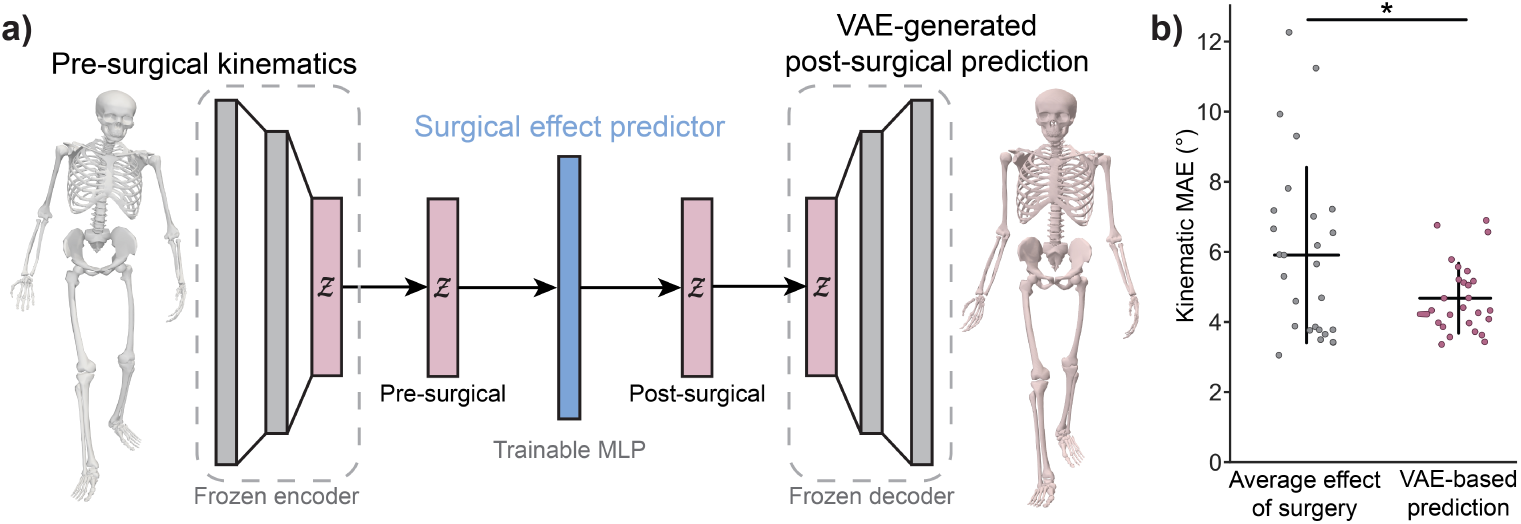
Generating predicted post-surgical kinematics from the GaitEncoder latent representation. **a)** We trained a small neural network (blue) to predict the post-surgical latent representation (*z*) from the pre-surgical one. The GaitEncoder encoder and decoder networks were frozen, and we evaluated the model on the generated post-surgical kinematics from held-out validation data. **b)** Post-surgical kinematics predicted by our VAE-based model were more accurate (*p=.004) than a baseline prediction that each individual’s kinematics improve in the same way (i.e., applying the mean change across the dataset).

### 2.5 Evaluating the final GaitEncoder model (n=657)

The evaluations previously described used a model trained on a subset of the dataset (n=381; 4 pathologies), to test performance on unseen movement-related conditions and individuals. We additionally train the final GaitEncoder model on all participants (n=657; 7 pathologies) and demonstrate low reconstruction errors (Supplemental Table S1) for various joints (MAE=2.9±1.3°) and translations (MAE=2.2±1.2 cm). Clinical diagnosis was not used during training, so to determine whether the latent space of the final model continues to be clinically meaningful, we trained support vector machines (SVMs) to classify controls ^41,42^ from individuals post-stroke ^42^, with Parkinson’s disease ^43^, and with hip osteoarthritis ^41^. Latent representations were averaged by participant, and SVMs were trained and evaluated using five-fold cross-validation. The balanced accuracy across 4 classes was 84.8% (chance is 25%).

## 3 Discussion

We present GaitEncoder, a generative foundation model that distills gait kinematics into a 16-dimensional latent representation and an interpretable scalar gait impairment score (DMU). To enable this, we curated a kinematically diverse dataset of 657 individuals (ages 8–86) walking at a comfortable speed, spanning seven pathologies and collected with multiple motion capture modalities. After training the model on a subset of this dataset, we evaluated it on four different downstream clinical tasks. We show that, after distilling kinematics into a 16-dimensional latent space, GaitEncoder can accurately reconstruct kinematics (MAE=3.5°) across diverse gait pathologies. Next, we found that the latent representation was sensitive to longitudinal improvements in gait that occurred during post-stroke recovery. The latent representation, coupled with the model’s generative capability, also enabled prediction of personalized responses to total hip arthroplasty. Additionally, GaitEncoder generalized to three disorders not seen during training and across multiple motion capture modalities. The model performed downstream tasks in a zero-shot setting on these unseen clinical populations. Fine-tuning on the held-out datasets further improved accuracy. This demonstrates that our model generalizes to unseen pathologies, could be readily applied in the clinic, and can be fine-tuned with a small amount of pathology-specific data to improve performance on specific downstream clinical tasks.

Unlike most machine learning models in biomechanics that are trained on data from a single pathology, GaitEncoder is trained on a dataset spanning seven different pathologies, enabling generalization to varied clinical tasks across populations both within and outside its training distribution. For example, GaitEncoder predicts disease severity scores in Parkinson’s disease, a condition withheld from training. It captures rehabilitation progress in a post-stroke patient measured with markerless motion capture, even though post-stroke gait was only represented by marker-based data during training. It also differentiates myotonic dystrophy and FSHD from controls using smartphone video–based kinematics, despite these neuromuscular diseases being held out from the training data. Together, these results show that the GaitEncoder latent space remains useful across motion capture modalities and for both seen and unseen pathologies. This can accelerate the development of models for a variety of gait-related conditions, with particular benefit in rare conditions, where collecting a large dataset is difficult.

Foundation models have proven useful in computer vision, animation, and modeling the physics of human movement in healthy populations; here, we extend this paradigm to clinical gait analysis. In the computer vision community, models can generate and infer realistic human movement from video or text, demonstrating strong performance for activities with unimpaired populations ^44,45^. In biomechanics, a recent model trained with a dataset of 270 individuals (99% unimpaired) has proven useful in estimating unmeasured quantities from (e.g., ground forces) from kinematic measurements ^29^, reducing the burden of capturing biomechanics data outside a laboratory setting. Variational autoencoders applied to joint kinematics have been used for disease detection ^34^ and for tokenizing movement as input to language models ^35^, but have yet to show broad generalizability. Leveraging a large (n=657), pathologically diverse dataset, we trained a foundation model that extends to clinical gait analysis, and we demonstrated that it generalizes across heterogeneous impairments and supports multiple clinically relevant downstream tasks.

GaitEncoder supports gait assessment across increasing levels of analytical complexity. The simplest output is the DMU, an interpretable score summarizing whole-body gait impairment. The DMU was inspired by the Gait Deviation Index ^1^, which applies a linear decomposition of lower-limb kinematics, but we extend this framework to whole-body kinematics using nonlinear dimensionality reduction. We demonstrate that the DMU differentiates individuals with neuromuscular diseases from control, whereas the linear approach used to create the Gait Deviation Index, applied to our whole-body kinematics dataset did not. This contrast suggests that incorporating nonlinearity, along with training on pathologically diverse data, may improve generalization. We also demonstrate that the DMU predicts commonly used clinical impairment scores for Parkinson’s disease and is sensitive to longitudinal changes from post-stroke rehabilitation (n=1). The data for myotonic dystrophy, FSHD, and the post-stroke evaluations were collected using smartphone videos in a gymnasium, physical therapy clinic, and in the home, highlighting the clinical feasibility of computing the DMU. We deployed the DMU computation to the cloud-based OpenCap platform so it can be automatically computed after a smartphone video–based gait analysis.

The latent representation itself enables a variety of more complex analyses. First, it represents a salient, automatically selected, and small set of features that can be used as input to models for a variety of downstream tasks. For example, models using hand-engineered gait features designed alongside clinicians ^17^ did not accurately differentiate myotonic dystrophy or FSHD from controls (44–53% accuracy). GaitEncoder features outperformed hand-engineered features in a zero-shot evaluation (62–68% accuracy), and accuracy further improved after fine-tuning on the pathology-specific datasets (75–81%). Second, the decoder can generate realistic gait trajectories that could result from hypothetical treatments. We trained a small surgical effect predictor model to map pre- to post-surgical latent gait representations for total hip arthroplasty, then used the decoder to generate predicted post-surgical kinematics. This personalized post-surgical gait pattern was more accurate than the current standard of assuming a population-average improvement. As commonly performed procedures such as hip replacement has been overutilized ^46^, the ability to identify patients who are unlikely to experience meaningful gait improvements may promote alternative or pre-surgical interventions. Although our improvement of 1.2° is modest, pre-surgical kinematics alone likely capture only a subset of the factors that determine post-surgical outcomes in this cohort. Intervention prediction models like this become especially powerful for conditions with many possible interventions, like cerebral palsy. The ability to visualize post-surgical gait predictions for various candidate interventions based on models that use demographics and baseline gait could be a powerful decision-support tool for both clinicians and families ^4^.

GaitEncoder’s latent representation could also inform precision rehabilitation. While gait speed is a widely used gait outcome, it cannot distinguish true recovery from compensation after stroke ^47^. Combining the DMU, a measure of kinematic normalcy, with a measure of task performance (e.g., gait speed) can identify which aspects of gait have recovered and where deficits remain ^48^. In our post-stroke case study, the patient’s DMU decreased rapidly before plateauing with a small amount of deviation from normative gait kinematics, while gait speed continued to improve slowly for four more weeks until it exceeded the normative average gait speed. At the time that gait speed had plateaued but DMU remained elevated, we used DMU’s interpretability to pinpoint the joints contributing most to the residual deficit, which could help guide targeted rehabilitation. Both DMU and intervention efficacy prediction models based on the GaitEncoder latent representation could support more personalized rehabilitation for a variety of neuro-musculoskeletal conditions.

These models could address the lack of sensitive functional outcome measures for neuromuscular disorders ^36^, which is hindering the development of therapeutics. For example, the control arm of a recent phase III clinical trial for FSHD showed no progression over 48 months ^49^, making it difficult to capture potential disease-modifying effects in the treatment arm. Latent representations and the DMU quantify changes in gait kinematics, providing a holistic characterization of movement quality, without requiring specific hand-engineered kinematic features (e.g., peak knee angle) ^17,34^. The DMU can detect aberrant movement patterns at any joint and time during the gait cycle, making it sensitive to the heterogeneous patterns of weakness and movement that are common in these conditions. Here, we show that DMU measured with smartphone videos can discriminate individuals with FSHD and myotonic dystrophy from controls better than hand-engineered features, which includes the clinical-standard gait speed. Furthermore, the DMU was sensitive to longitudinal changes in gait in our post-stroke case study. This supports the potential of GaitEncoder’s latent gait representation to holistically capture the quality of the complex task of walking, and to detect changes in movement quality from natural disease progression and treatment.

It is important to acknowledge the limitations of our study. First, although Gait-Encoder generalizes across clinical populations and downstream tasks, it was trained and evaluated on walking data and likely does not directly generalize to other activities. Next, our model can encode and reconstruct full body motion or for a subset of joints but is only trained using a specific biomechanical model ^50^; however, this skeletal model is commonly used in open-source biomechanics tools, like OpenCap ^18^ and AddBiomechanics ^51^. Additionally, we used marker-based and video-based kinematics here but did not test the model using IMU-based kinematics. Furthermore, our model only considers kinematic data, but some disorders such as knee osteoarthritis may benefit more from latent representations that include dynamics ^29^.

GaitEncoder provides a compact representation of gait that generalizes across various unseen pathologies, downstream clinical tasks, and motion capture modalities. In combination with recent advances in mobile sensing, it provides a modern machine learning framework for converting salient information about movement into actionable clinical insights. We open-source GaitEncoder and deploy it in the cloud, enabling automated DMU scoring, latent feature extraction, and straightforward fine-tuning for novel downstream tasks. This work demonstrates how clinically oriented foundation models of human movement can facilitate both personalized and large-scale movement analysis to improve the treatment for a wide range of movement-related conditions.

## 4 Methods

### 4.1 Dataset

We aggregated eight gait datasets ^17,41–43,52–55^ (Table 1) representing individuals with and without movement-related diagnoses, spanning seven conditions. We included six datasets with marker-based motion capture and two with markerless motion capture. The training set included post-stroke, knee osteoarthritis, hip osteoarthritis, cerebral palsy, and control cohorts. We withheld Parkinson’s disease, myotonic dystrophy, FSHD, post-stroke, and some controls entirely for evaluation of out-of-distribution generalization (Figure 2a). All walking trials were recorded at a comfortable or self-selected pace without assistive devices, and each dataset provided either raw markers or OpenSim-processed kinematics ^56,57^ using the Rajagopal musculoskeletal model ^50^.

**Table 1.**
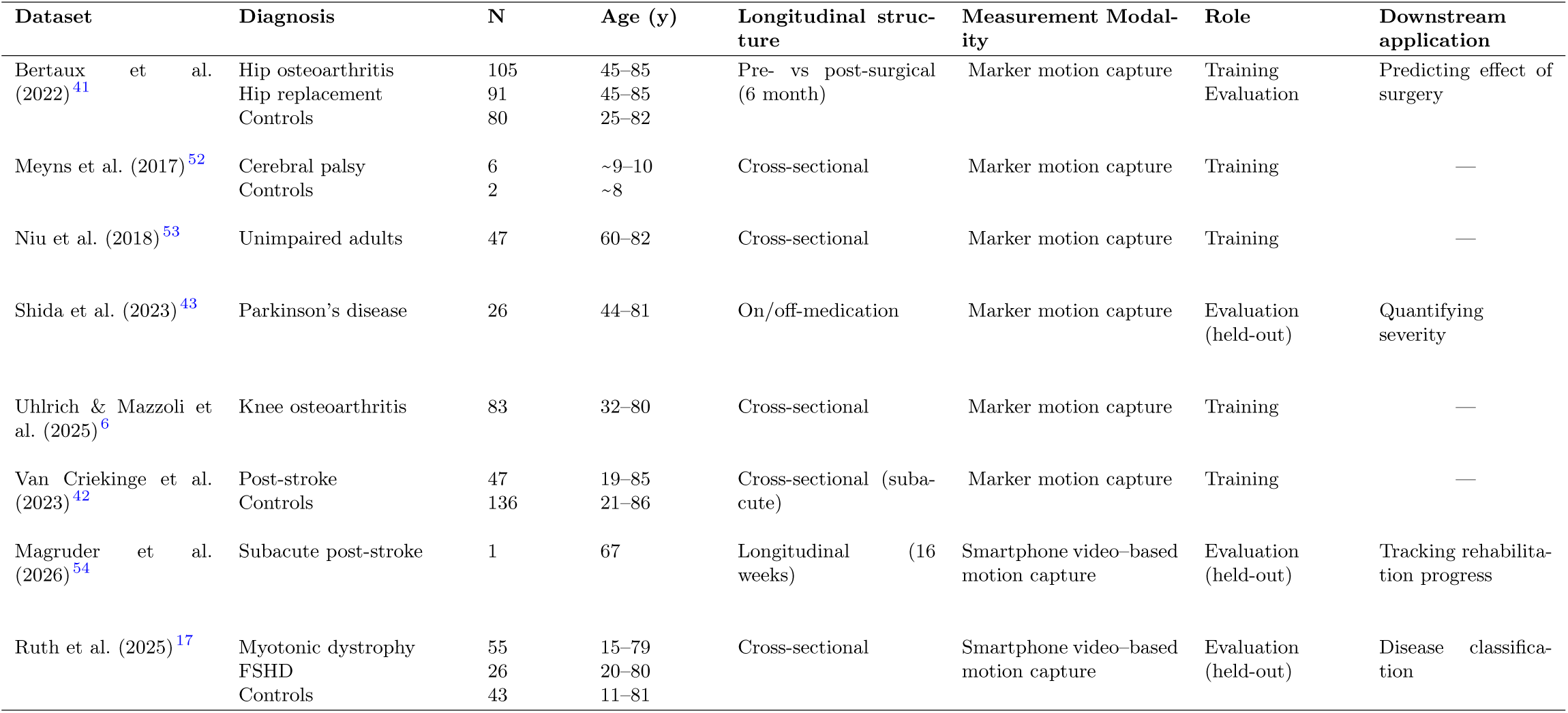
Summary of datasets (total n=657) used to train and evaluate the GaitEncoder model. All datasets consist of overground walking at a comfortable or self-selected pace without assistive devices. Marker-based datasets were processed to joint kinematics using OpenSim, while smartphone-based datasets used OpenCap. Datasets designated as “held-out” were not included during model training or validation and were used exclusively to assess out-of-distribution generalization and clinical relevance.

For the marker-based datasets, we rotated markers such that each person walked in the positive x-direction and used AddBiomechanics ^51^ with the Rajagopal model ^50^ to compute kinematics. We processed video-based datasets using OpenCap (neuromuscular disease dataset) ^18^ or Model Health Inc. (post-stroke case study) and the same biomechanical model. We segmented kinematics into gait cycles using foot marker positions; when a patient had known lateralized impairment, only the segmentation beginning with their more impaired side was included. Left leg strides were flipped to mimic right leg motion to avoid encoding laterality. We computed gait speed using pelvis translation in the x direction over the stride and stride time.

All data on individuals with Parkinson’s disease (n=26), myotonic dystrophy (n=55), and FSHD (n=26) were withheld entirely from training, and the remaining datasets were split 70/30 by participant into training and validation (Figure 2a). For the unimpaired OpenCap cohort, 70% were used in training while the remaining 30% were held out entirely for downstream evaluation. The longitudinal patient recovering from stroke was held out from training/validation, but other post-stroke data were used in model training. Kinematics from unimpaired individuals were used to compute joint-wise means and standard deviations, which were there used to standardize all kinematic inputs to the VAE.

### 4.2 VAE Training

We used the aggregated dataset to train a VAE to distill 32 joints by 24 stride-normalized time points into a 16-dimensional latent representation of gait. The model comprised an encoder, decoder and regression head. The encoder compressed the flattened input joint vector (768 features) through 2 fully-connected layers (256 and 128) then linearly to the 16-dimensional latent space. The decoder reconstructed the original input vector with inverted architecture (128 then 256 features). The regressor head used a learnable linear layer to predict the gait speed by stride from the latent space. The loss function had 3 terms: reconstruction, Kullback-Liebler Divergence, and gait speed error. Reconstruction was penalized by MAE across all joints. Kullback-Liebler divergence assumes a prior normal distribution of the latent space which promotes a continuous latent manifold ^31^. The linear layer from the latent space predicted gait speed to weakly supervise the model, ensuring the latent space is linearly interpretable for a common clinically used metric. We deliberately avoided using explicit labels such as diagnosis during training, since this could hinder the encoding of unseen disorders. Instead, gait speed was chosen because it is a continuous, clinically meaningful measure of mobility across a wide range of conditions ^58,59^. We performed a hyperparameter search for various latent sizes (4 to 40 dimensions; Figure S1), each trained for 400 epochs with early stopping. We selected a latent size of 16-dimensions based on the elbow point of reconstruction MAE and minimal features. For interpretability, reconstruction MAE is reported in degrees despite inclusion of stride time (s) and pelvis translations (m) in the feature set. We also found that a transformer layer prior to the fully connected layers did not improve reconstruction error.

We further distilled the 16-dimensional latent space into a scalar impairment value based on kinematic deviations from normative embeddings. Our aggregated dataset of unimpaired individuals includes a large age range, with the inclusion criterion of studies being highly variable. Thus, we chose a subset of all unimpaired persons as a high-performing cohort to be those between 18 and 65 years of age with a gait speed greater than or equal to 1.2 m/s, since gait speed declines with age ^60^ and is a predictor of cognitive and physical impairment ^61^. We aggregated the latent representation for each high-performing individual and computed the deviation from this distribution using Mahalanobis distance ^1^ which accounts for the variance within each feature as the deviation from mean unimpaired (DMU) as a scalar impairment score.

We make multiple versions of the VAE publicly available. Unless otherwise noted, the model used in this paper was trained on a set of four gait pathologies (n=381, described in Methods: Dataset). In our Github repository, we also provide the Gait-Encoder fine-tuned for FSHD, for myotonic dystrophy, and the model trained on all seven gait pathologies and all participants (n=657). The DMU that is automatically computed on the OpenCap platform uses this model trained on the entire dataset, and we recommend using this version of the model in future applications as it is trained on the most diverse data (Supplemental Table 1).

### 4.3 Downstream application 1: Identification of rare neuromuscular diseases using gait kinematics

We calculated the DMU from GaitEncoder in a zero-shot setting to determine whether the model could differentiate neuromuscular disorders from controls without pathology-specific training. The myotonic dystrophy (n=55), FSHD (n=26), and control (n=13) cohorts used for evaluation were held out of training. For comparison with the linear approach used for Gait Deviation Index ^1^, we also used single vector decomposition with 16 features and Mahalanobis distance for direct comparison. We performed one-sided t-tests (*α*=.05) with the a priori assumption that those with neuromuscular diseases will have higher scores than the control cohort for both our DMU method and the linear approach ^1^.

Additionally, we trained SVMs with and without fine-tuning to classify individuals with myotonic dystrophy from controls. We evaluated classification performance using 5-fold cross-validation. To fine-tune the VAE, we froze all but the layer connected to the latent layer for the encoder and decoder. We also added a new linear layer to predict classes from the latent space, and a new model was fine-tuned for each fold. The final fine-tuned models for each fold were used to train SVMs as before. Thus, for each fold, the held-out participants were not included in either VAE fine-tuning or SVM training. We compared the performance of latent-based SVM to SVMs using hand-engineered features described in Ruth et al. (2025) for walking and across 9 activities ^17^. This was performed for each neuromuscular disease against the OpenCap control cohort. We used McNemar’s test of proportions to determine which SVMs outperformed hand-engineered gait features. Final fine-tuned models for FSHD and myotonic dystrophy were trained with all patients from these cohorts and the OpenCap control. DMU was recomputed using the fine-tuned latent space and OpenCap training control cohort. We performed one-sided t-tests as previously described for the DMU scores from fine-tuned VAEs between control and either FSHD or myotonic dystrophy.

### 4.4 Downstream application 2: Regressing patient impairment with clinician and self-reported scales

We used linear regression and leave-one-subject-out cross-validation to predict self-reported (UDPRS-II) and clinician-rated (UPDRS-III) Parkinson’s disease severity scores from the DMU and latent representations of gait. The 26 individuals with Parkinson’s disease were held out from VAE training. On- and off-medication states were treated as separate data points in subsequent analyses. We report p-values corrected for multiple comparisons using the Benjamini-Hochberg procedure to control for the false discovery rate ^62^.

### 4.5 Downstream application 3: Tracking patient recovery during subacute, post-stroke rehabilitation

We collected comfortable walking trials using the OpenCap markerless motion capture system during at-home data collection sessions with two smartphone videos longitudinally for 16 weeks post-stroke (n=1). The participant provided informed consent to a protocol approved by the Institutional Review Board at the University of Utah, including the public sharing of data. Kinematics were computed from videos using Model Health (Salt Lake City, Utah, USA) and the Rajagopal model^50^. Due to day-to-day variability during rehabilitation, we used a causal, moving average of 3 weeks with ramp-up to smooth all features and simulate the data availability when monitoring recovery in the clinic. This was a case study, so no statistical testing was performed, although common clinically measured gait features for stroke recovery—gait speed and step length symmetry—were also extracted as clinical comparators. These features were computed using the open-source OpenCap gait analysis tool ^18^.

At 16 weeks post-stroke, both gait speed and DMU were stable, but DMU remained elevated and, visually, mild hemiparesis was noticeable (Supplemental Video S2) in comparison to the average of unimpaired gait. We quantified this by observing the change in DMU score as each joint was replaced with the average unimpaired kinematics for that joint. Each joint was substituted independently, and the resultant change in computed DMU was recorded. The four joints to impact DMU the most were evaluated qualitatively to determine whether these joints showed worsening consistent with post-stroke gait patterns.

### 4.6 Downstream application 4: Predicting kinematic changes after total hip arthroplasty

We trained a feedforward neural network with a single hidden layer (64 nodes) to model the change in latent representation of gait after total hip replacement. Our dataset included gait data from those with hip osteoarthritis (n=105) and those who returned (n=91) for a second session 6 months after total hip arthroplasty surgery. For a baseline model comparison, we computed the change in stride-level kinematics due to surgery as the average estimated effect of surgery (Eq. 1). This change was computed using the 70% of individuals included in the training set (Eq. 2). During training, patients before and after surgery were paired, so no person would be in training and validation for different timepoints. The average estimated change was applied to the pre-surgical kinematics of the validation set and the MAE of the lower limbs was computed (Eq. 3), where q is an individual’s kinematics from the training set, j, or validation set, i.

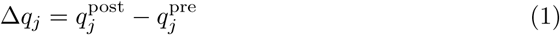

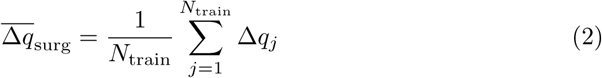

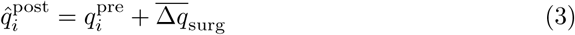

The surgical effect predictor was trained using the same training set used to construct the average surgery effect model; we did not use the validation set for hyperparameter tuning or early stopping. The input to the surgical effect predictor is the pre-surgical GaitEncoder latent representation (Figure 6), which was not fine-tuned (frozen) for this application. We averaged the latent representation for multiple gait cycles for each individual, so each individual’s training data comprised one pre-surgical and one post-surgical latent representation. The output of the model is the post-surgical latent, and the loss function minimized Euclidean distance between predicted and ground truth post-surgical latent vector. After training, we generated post-surgical kinematics using the frozen GaitEncoder decoder. We compared the VAE-based method to the average effect of surgery on the validation set using MAE of kinematic timeseries with a two-sided, paired t-test (*α*=.05).

## Supporting information

final_weeks_sidebyside.mp4

week16_overlay.mp4

## Declarations

### Data availability

All processed kinematics data, homogenized around the Rajagopal model, used in model training and evaluation are publicly available on SimTK (https://simtk.org/projects/gaitencoderdata) ^54^.

### Code availability

The trained models and source code is freely available on GitHub (https://github.com/rdmagruder/GaitEncoder) for research. We also integrate GaitEncoder in OpenCap gait analysis (https://github.com/opencap-org/opencap-processing) and automatically compute DMU on the OpenCap platform (https://www.opencap.ai/). Intuitive visualization and stride generation can be found on our project page (https://huggingface.co/spaces/rdmagruder/gaitencoderprojectpage).

### Author contribution

RDM and SDU provided conceptualization, methodology, and initial drafting. RDM, SG, and AF worked on data curation and analysis. All authors critically revised the manuscript and approved the final version.

