## Supplementary Appendix for "GaitEncoder: A Foundation Model of Gait Kinematics for Diverse Clinical Applications and Pathologies"

#### Latent Space Dimensionality

To determine the smallest possible latent space size required to accurately embed gait kinematics, we trained VAEs with latent sizes ranging from 4 to 40 (Figure S1a). Models were trained using the same training validation splits and evaluated with mean absolute reconstruction error of the validation set (n=155, 4 pathologies). We swept for hyperparameters (learning rate, number of layers, layer dimensions, loss weighting, dropout, masking) at each latent size and recorded the best performing model. Reconstruction improved with increasing latent size; however, we chose 16 dimensions because this was the clear inflection point, and a smaller latent will likely generalize better and serve as a better input to downstream models. Using a VAE with a 16-dimensional latent, we included a transformer head prior to the VAE, and swept for hyperparameters. The inclusion of a transformer layer did not improve reconstruction error, so the VAE with MLP encoder and decoder was chosen as the final model (Figure S1a). All subsequent evaluations were performed using this model architecture. All persons in the validation set were used for selecting the best model, however notably unimpaired persons have lower reconstruction errors than the full validation cohort (Figure S1b).

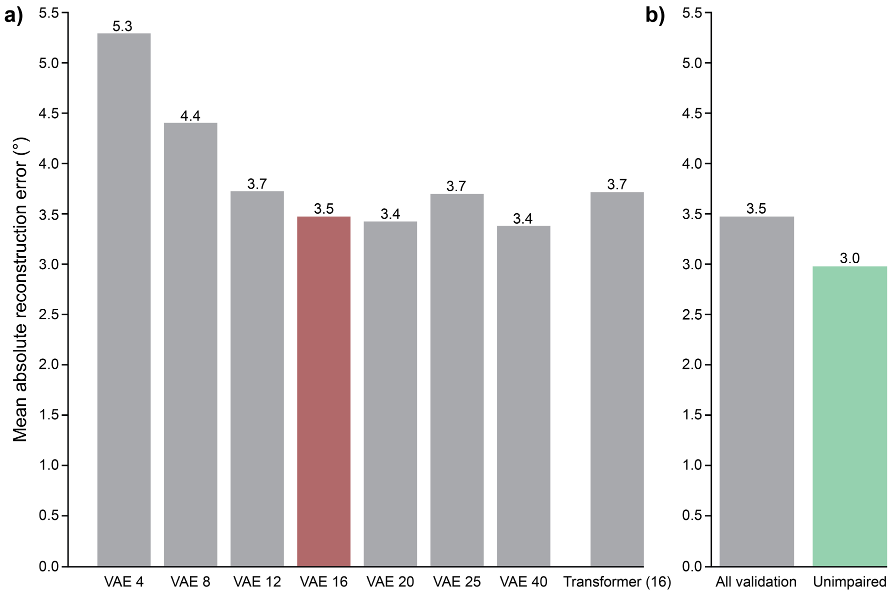


**Fig. S1 A latent space with 16 features is sufficient to represent gait.** a) Reconstruction error across all joints for the validation dataset. We selected a 16-dimensional latent space as it minimized both reconstruction error and latent dimensionality. We tested adding a transformer head prior to the MLP variational autoencoder (VAE) layers, but it did not improve reconstruction error over the MLP-only VAE. b) Reconstruction error for unimpaired persons is smaller (3.0°) than across all persons (3.5°) in the validation set.

### Parkinson's Disease Scoring on vs. Off Medication

Levodopa is a common dopamine replacement therapy for Parkinson's disease and can improve gait kinematics. The Parkinson's disease dataset[1] comprised both on- and off-medication states. We performed an exploratory analysis of whether models predicting UPDRS scores from GaitEncoder had different accuracy for on- and off-medication. The strength of correlation between the DMU and UPDRS-III was similar when using on-medication, off-medication, and an aggregated dataset: r=0.54, 0.49, and 0.51, respectively. Thus, we used the aggregated dataset and leave-one-subject-out analysis in the main analysis of this dataset (Figure 4).

#### Stroke Recovery Case Study Video

We collected smartphone video–based kinematics of a person recovering longitudinally post-stroke. The participant was held out of training, and we used the GaitEncoder to generate the impairment score over time. Supplemental Video S1 shows the measured kinematics (grey) and the reconstructed kinematics (blue). Strides shown are presented sequentially (weeks 5, 7, 10, and 16 post-stroke) to illustrate the patient's recovery trajectory. Supplemental Video S2 overlays average kinematic curves (grey) with the participant at week 16 (pink). While walking at a normative speed, DMU still has not reached 1.0, and there are some visible kinematic asymmetries.

**Video S1.** See `final_weeks_sidebyside.mp4`

**Video S2.** See `week16_overlay.mp4`

#### MAE of the GaitEncoder Model (Trained on n=657)

Supplemental Table S1 reports the mean absolute error (MAE) of the final GaitEncoder model (trained on n=657) for various translations and joint angles.

| | All Individuals (n=657) | | Unimpaired (n=308) | | Impaired (n=349) | |
|---|---|---|---|---|---|---|
| **Joint** | **Mean ± SD** | **Median** | **Mean ± SD** | **Median** | **Mean ± SD** | **Median** |
| **Translations (cm)** | | | | | | |
| Pelvis Tx (forward) | 4.4 ± 3.0 | 3.6 [2.2, 5.6] | 3.8 ± 2.8 | 3.3 [1.8, 5.0] | 4.8 ± 3.1 | 3.8 [2.6, 6.4] |
| Pelvis Ty (lateral) | 0.7 ± 0.5 | 0.5 [0.4, 0.7] | 0.6 ± 0.4 | 0.5 [0.4, 0.7] | 0.7 ± 0.5 | 0.5 [0.4, 0.8] |
| Pelvis Tz (vertical) | 1.6 ± 0.9 | 1.4 [1.0, 1.8] | 1.4 ± 0.7 | 1.2 [1.0, 1.6] | 1.7 ± 0.9 | 1.5 [1.1, 2.1] |
| **Whole-Body Joints (°)** | | | | | | |
| All joints | 2.9 ± 1.3 | 2.6 [2.3, 3.2] | 2.7 ± 1.5 | 2.5 [2.2, 2.9] | 3.0 ± 1.2 | 2.8 [2.4, 3.5] |
| Lower limb joints | 2.5 ± 0.9 | 2.4 [2.0, 2.9] | 2.3 ± 0.6 | 2.2 [1.8, 2.5] | 2.8 ± 1.0 | 2.6 [2.2, 3.2] |
| **Individual Joints (°)** | | | | | | |
| Pelvis tilt | 1.5 ± 0.6 | 1.3 [1.0, 1.8] | 1.3 ± 0.4 | 1.2 [1.0, 1.5] | 1.6 ± 0.7 | 1.5 [1.2, 2.0] |
| Pelvis list | 1.4 ± 0.6 | 1.3 [1.0, 1.7] | 1.3 ± 0.5 | 1.1 [0.9, 1.5] | 1.5 ± 0.6 | 1.4 [1.1, 1.8] |
| Pelvis rotation | 2.3 ± 1.2 | 2.0 [1.6, 2.7] | 2.0 ± 0.9 | 1.9 [1.5, 2.4] | 2.6 ± 1.4 | 2.2 [1.7, 3.0] |
| Lumbar extension | 1.9 ± 1.0 | 1.7 [1.3, 2.3] | 1.7 ± 0.8 | 1.6 [1.2, 2.1] | 2.1 ± 1.2 | 1.8 [1.3, 2.6] |
| Lumbar bending | 2.1 ± 1.0 | 1.9 [1.5, 2.4] | 1.9 ± 0.8 | 1.8 [1.3, 2.3] | 2.3 ± 1.1 | 2.0 [1.6, 2.7] |
| Lumbar rotation | 2.7 ± 2.1 | 2.3 [1.7, 3.1] | 2.6 ± 1.9 | 2.2 [1.7, 3.1] | 2.8 ± 2.3 | 2.3 [1.8, 3.2] |
| Hip flexion | 2.6 ± 1.1 | 2.3 [1.9, 3.1] | 2.2 ± 0.7 | 2.1 [1.7, 2.5] | 3.0 ± 1.2 | 2.8 [2.1, 3.6] |
| Hip adduction | 1.9 ± 0.8 | 1.8 [1.4, 2.3] | 1.7 ± 0.7 | 1.6 [1.3, 2.0] | 2.1 ± 0.9 | 1.9 [1.6, 2.4] |
| Hip rotation | 2.9 ± 1.1 | 2.7 [2.2, 3.3] | 2.7 ± 0.8 | 2.6 [2.1, 3.1] | 3.0 ± 1.2 | 2.8 [2.4, 3.4] |
| Knee angle | 3.5 ± 1.5 | 3.3 [2.5, 4.1] | 3.0 ± 1.1 | 2.9 [2.3, 3.5] | 4.0 ± 1.7 | 3.6 [2.9, 4.8] |
| Ankle plantarflexion | 3.5 ± 2.1 | 2.9 [2.2, 4.0] | 3.1 ± 1.6 | 2.7 [2.1, 3.5] | 3.8 ± 2.4 | 3.3 [2.4, 4.3] |

**Table S1 Reconstruction error of the final model.** Reconstruction mean absolute error (MAE) was computed for each joint and translation of each participant. Values are reported as mean ± standard deviation and median [Q1, Q3] across the average of an individual. Lower-limb joints include the lumbar spine (3 DoF), pelvis rotations (3 DoF), hip joints (3 DoF), knee flexion (1 DoF), and ankle plantarflexion (1 DoF). All joints additionally include upper-extremity joint angles and the subtalar joint.
